# Causal empirical estimates suggest COVID-19 transmission rates are highly seasonal

**DOI:** 10.1101/2020.03.26.20044420

**Authors:** Tamma Carleton, Kyle C. Meng

**Affiliations:** University of Chicago, Dept. of Economics and Energy Policy Institute; Bren School of Environmental Science and Management, Dept. of Economics, and Environmental Markets Solutions Lab (emLab), UC Santa Barbara and National Bureau of Economic Research

**Author notes:** Equal contributions. We thank our families for their patience with this project during a trying period. We received helpful comments and feedback from Jonathan Dingel, Antony Millner, Ishan Nath, Jonathan Proctor, and Ashwin Rode. Jules Cornetet and Shopnavo Biswas provided excellent research assistance. The authors declare no conflicts of interest. All data and replication files can be found at https://github.com/emlab-ucsb/COVID-seasonality.

## Abstract

Nearly every country is now combating the 2019 novel coronavirus (COVID-19). It has been hypothesized that if COVID-19 exhibits seasonality, changing temperatures in the coming months will shift transmission patterns around the world. Such projections, however, require an estimate of the relationship between COVID-19 and temperature at a global scale, and one that isolates the role of temperature from confounding factors, such as public health capacity. This paper provides the first plausibly causal estimates of the relationship between COVID-19 transmission and local temperature using a global sample comprising of 166,686 confirmed new COVID-19 cases from 134 countries from January 22, 2020 to March 15, 2020. We find robust statistical evidence that a 1°C increase in local temperature reduces transmission by 13% [−21%, −4%, 95%CI]. In contrast, we do not find that specific humidity or precipitation influence transmission. Our statistical approach separates effects of climate variation on COVID-19 transmission from other potentially correlated factors, such as differences in public health responses across countries and heterogeneous population densities. Using constructions of expected seasonal temperatures, we project that changing temperatures between March 2020 and July 2020 will cause COVID-19 transmission to fall by 43% on average for Northern Hemisphere countries and to rise by 71% on average for Southern Hemisphere countries. However, these patterns reverse as the boreal winter approaches, with seasonal temperatures in January 2021 increasing average COVID-19 transmission by 59% relative to March 2020 in northern countries and lowering transmission by 2% in southern countries. These findings suggest that Southern Hemisphere countries should expect greater transmission in the coming months. Moreover, Northern Hemisphere countries face a crucial window of opportunity: if contagion-containing policy interventions can dramatically reduce COVID-19 cases with the aid of the approaching warmer months, it may be possible to avoid a second wave of COVID-19 next winter.

## Introduction

In late 2019, a novel virus strain from the family *Coronaviridae*, referred to as 2019-nCoV (or SARS-CoV-2) began spreading throughout China.^1^ Central among 2019-nCoV concerns are its high transmissivity, potentially severe flu-like symptoms, and high case fatality rates.^2^ In the ensuing months, the virus has transmitted globally, prompting the World Health Organization to declare a pandemic on March 11, 2020. At the time of this writing, cases of COVID-19, the disease caused by 2019-nCoV, have been detected in 134 countries (Fig. 1A).

**Figure 1:**
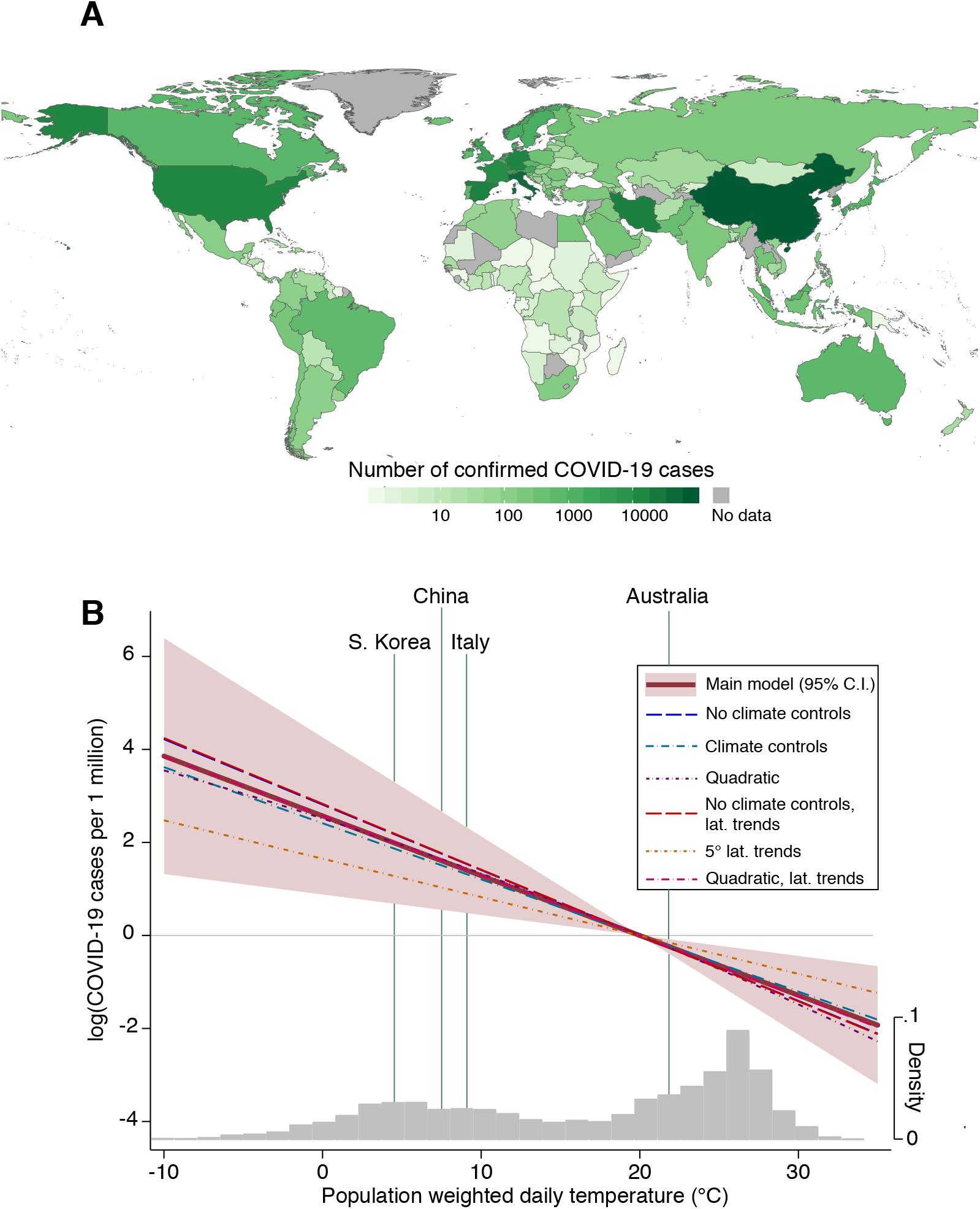
Empirical estimates of a global COVID-19 and local temperature relationship. Panel A shows total confirmed cases of COVID-19 across 134 countries, as of March 15, 2020. Darker colors indicate a higher total number of cases; grey indicates that no data are available. Panel B shows the estimated response of new COVID-19 cases to daily, population-weighted average temperature across seven distinct statistical models. The *y*-axis is in units of the natural log of total new cases per 1 million population. Our benchmark specification is indicated by the solid red line (col. 5, Table S1), representing a 13% decline in the case rate per 1°C increase in temperature, with a 95% confidence interval of [−21%, −4%] shaded in red. Coefficients and model details for all specifications are shown in Table S1: col. 1 (no climate controls), col. 2 (climate controls), col. 3 (quadratic), col. 4 (no climate controls, 25°C latitudinal band trends), col. 6 (quadratic, 25°C latitudinal band trends), and col. 7 (5°C latitudinal band trends). The grey histogram reflects the distribution of population-weighted average daily temperature in °C across all countries in the sample between January 22, 2020 and March 15, 2020. Average temperatures for select countries are indicated by green vertical lines.

Much remains unknown about COVID-19. An important question concerns the conditions that affect transmission. Previous studies show that transmission of H3N2, 2009 H1N1, and other strains of influenza are sensitive to environmental conditions, and in particular decline with higher temperatures and humidity.^3–7^ If COVID-19 exhibits similar climatic sensitivities, it is possible that warmer temperatures during the boreal summer months may lower transmission rates for Northern Hemisphere countries while cooler temperatures increase transmission rates for Southern Hemisphere countries over the same period. Such climatic determinants of transmission present opportunities and challenges for policymakers and scientists, as seasonal changes both open windows of opportunity to constrain the spread of the virus, while also accelerating rates of transmission in locations where stringent public health measures have not yet been put in place.

Several research efforts are underway to understand the temperature-COVID-19 transmission pattern. However, to date, results from existing studies compare temperature and COVID-19 cases across countries or across regions within individual countries. These empirical frameworks conflate location-specific characteristics – such as testing capacity, population density, and health services – with temperature variation.^8–10^ Because temperature is correlated with many (often unobservable) confounding factors, such cross-sectional comparisons may not have a causal interpretation.^11–13^ For example, countries that are cooler on average also tend to have higher income per capita^14^ which may affect the number of new COVID-19 cases by enabling more public health measures such as testing and hospitalizations. Moreover, to our knowledge, prior empirical estimates of the COVID-19-temperature relationship only examine cases within China, which may not be suitable for understanding the temperature-COVID-19 relationship globally. Additionally, several studies compare temperature and COVID-19 transmission across countries, but make no attempt to estimate a statistical relationship between temperature and COVID-19 cases^15,16^ or to generate empirically-based seasonal projections.

In this paper we statistically estimate a plausibly causal, global relationship between temperature and COVID-19 transmission and use these empirical estimates to generate high-resolution seasonal projections of new COVID-19 cases under the changing global climate over the next 11 months. To achieve this, we collect daily temperature, precipitation and specific humidity for every 0.25° latitude by 0.25° longitude pixel of the planet generated by a state-of-the-art climate reanalysis model in near real-time. We aggregate pixel-level weather variables to the country level using population weights and link to daily country-level reports of new COVID-19 cases across the world for the period between January 22, 2020 to March 15, 2020 (see Data section of Methods). This country-by-day global longitudinal dataset enables us to statistically estimate a global function between temperature and new cases of COVID-19 per 1 million people. In particular, we leverage the large existing climate econometrics literature^13^ to employ a statistical model containing a suite of semi-parametric controls designed to isolate the effect of temperature from potentially confounding factors that also influence new COVID-19 cases. These controls account for: time-invariant differences in population characteristics across countries, such as differential population densities and healthcare systems; temporal shocks that influence the pattern of global COVID-19 events, such as the WHO pandemic designation; and seasonal trends that vary latitudinally across the globe. Together, these controls aim to isolate idiosyncratic daily variations in temperatures experienced by the same population; we show that results are consistent across a range of alternative statistical models, each of which varies the stringency of these controls (see Statistical Model section of Methods).

## Results

We find robust statistical evidence that higher temperatures lower the number new cases of COVID-19 in a given population. In our main statistical specification, we find that a 1°C increase in local temperature reduces new COVID-19 cases per 1 million people by 13%, with a 95% confidence interval of [−21%, −4%] (Fig. 1B and col. 1 of Table S1). This result is robust to controlling for precipitation and specific humidity, neither of which exhibit statistically significant effects (although point estimates on humidity are negative, consistent with prior evidence for influenza^5^) (Fig. 1B and col. 2 of Table S1). Our findings are also robust to inclusion of a range of semi-parametric controls, including controlling for seasonal patterns of temperature that are common across countries within particular latitude bands of varying width (Fig. 1B and cols. 4-7 of Table S1). In our benchmark model (col. 5 of Table S1), we control for separate nonlinear trends for countries that fall within each 25° latitude band across the globe. This model accounts for spatially-varying trends in seasonal factors that may correlate both with COVID-19 cases and with temperature. Decreasing the width of these latitudinal bands, and thus imposing more restrictive controls on spatial patterns of seasonality, yields a coefficient statistically indistinguishable from that in our benchmark model (col. 7 of Table S1). These estimates are proportional, implying that the *level change* in the COVID-19 case rate per unit change in temperature will be higher in populations where the epidemic has already taken a strong hold. In contrast, lower temperatures will lead to far fewer individuals infected in a population with a low overall case rate. However, we detect no evidence of nonlinearity in this proportional response (Fig. 1B and col. 3 of Table S1); a 1°C increase in local temperature increases the case rate by 13% on average whether a country is presently relatively cold (e.g. South Korea), or relatively warm (e.g. Australia).

These estimates relate new COVID-19 cases to plausibly random daily variation in local temperatures. Thus, they reflect multiple possible channels through which changes in temperature can influence new cases. First, temperature is likely to have a biological effect on the virus 2019-nCoV itself, as has been documented in lab experiments for influenza.^3^ Second, temperature may change people’s behavior, and thus change the number of new cases caused by a single infected individual. For example, warming temperatures encourage outdoor activities,^17^ possibly changing the intensity of social interaction. Finally, temperature has a direct effect on other causes of morbidity and mortality, with particularly large impacts on elderly populations.^18–21^ Thus, temperature may increase susceptibility to COVID-19 by lowering overall health.

We combine the estimated temperature-COVID-19 relationship shown in Fig. 1B with expected seasonal temperatures over the next 11 months to project the effect that upcoming seasonal variation in temperature is likely to have on new cases of COVID-19 at high resolution across the globe. Fig. 2A shows, at a 0.25° latitude by 0.25° longitude resolution, the estimated percentage change in new COVID-19 cases around the globe caused by temperature changes between March 2020 and July 2020 (see Fig. S3 for 95% confidence interval). Fig. 2B collapses these projections to the country-level, plotting projected percentage changes for each country (*x*-axis) by latitude of each country’s area-weighted centroid (*y*-axis). The 20 countries with the highest current rate of COVID-19 cases per 1 million people are indicated with red dots; notably, the vast majority of the countries with high rates of COVID-19 cases lie above the equator, where temperatures are currently suppressed in the boreal winter. The projections shown in Figs. 2A and 2B reveal a clear hemisphere-dependent pattern in near-future temperature-driven COVID-19 transmission. By July 2020, we project, all things equal, that higher seasonal temperatures will drive COVID-19 transmission rates down by 43% relative to March 2020 on average across Northern Hemisphere countries. During that same period, transmission rates for Southern Hemisphere countries will rise by 71% on average. However, as Figs. 2C and 2D show, this relief for northern countries during the boreal summer is temporary. Expected cooling by January 2021 implies that average COVID-19 transmission is 59% higher than in March 2020 in northern countries. Over this same period, warming in southern countries warming lowers transmission by 2%.

**Figure 2:**
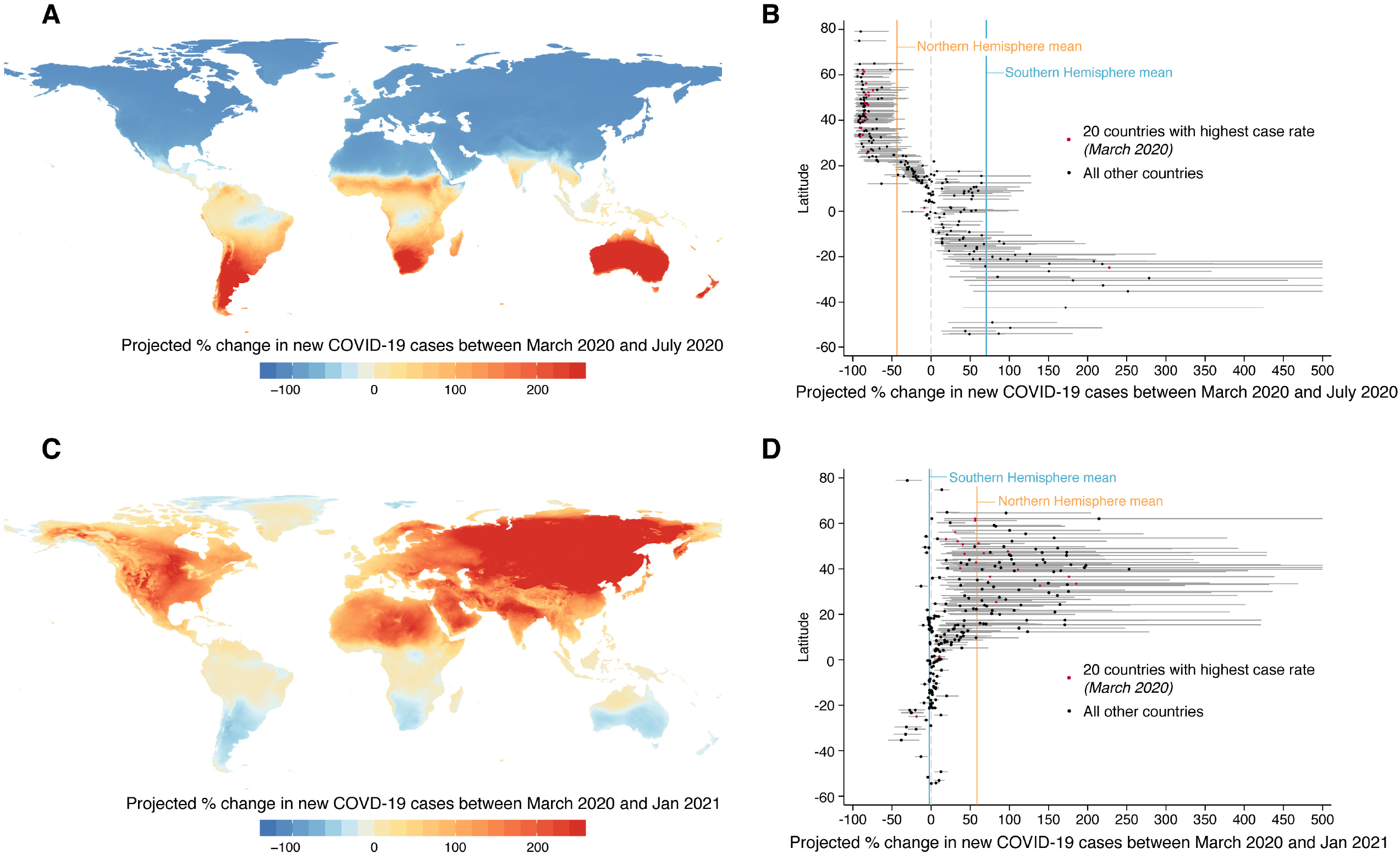
Spatial distribution of projected future COVID-2019 case rates due to changes in temperature. Panel A shows the projected percent change in new COVID-2019 cases per 1 million people due to changes in temperature between March 2020 and July 2020 at a 0.25° latitude by 0.25° longitude resolution. Panel B shows the same projected percentage change at the country-level, where countries are ordered vertically by latitude based on area-weighted centroids. Circles show projected change under mean estimates of the linear temperature-COVID-2019 effect, 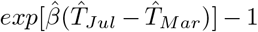 where 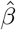 is from equation (1) and 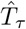 represents expected monthly temperature for month *τ*. Red circles indicate 20 countries with highest rate of COVID-19 cases per 1 million people, as of March 15, 2020. Grey error bars indicate 95% confidence intervals for each country; all confidence intervals are censored at 500%. Panel C (D) replicates Panel A (B) but for projected percentage changes between March 2020 and January 2021. These estimates reflect elevated or depressed transmission risk due only to temperature changes.

Fig. 3 demonstrates the temporal trajectory of these seasonal shifts in the global pattern of COVID-19. Each line represents a time series of seasonal projections of new cases of COVID-19 per 1 million individuals for each country and every month between April 2020 and January 2021; Northern Hemisphere countries are in orange and Southern hemisphere countries are in blue, with solid lines indicating hemisphere average effects. While temperatures in coming months are likely to depress new COVID-19 cases in northern countries, these gains are counterbalanced by large increases in risk for southern countries and a dramatic spatial reversal in the case burden by fall of 2020.

**Figure 3:**
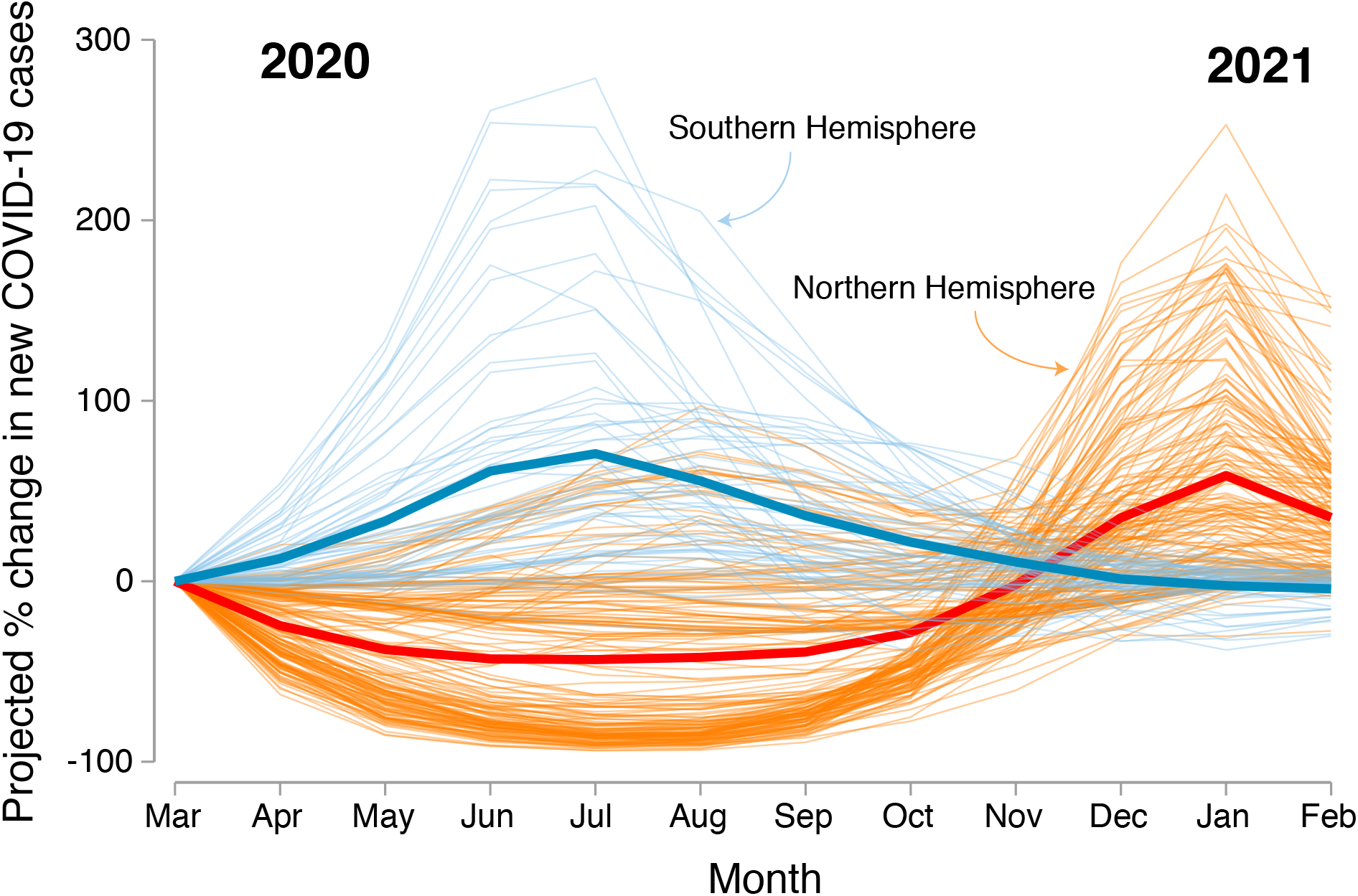
Time series of projected new COVID-2019 cases due to changes in temperature. Each thin line shows a country’s projected percent change in new COVID-2019 cases per 1 million people due to changes in temperature between March 2020 and the subsequent 11 months. Projections are generated under mean estimates of the linear temperature-COVID-2019 effect, 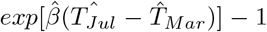, where 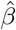 is from equation (1) and 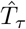 represents expected monthly temperature for month *τ*. These estimates reflect elevated or depressed transmission risk due only to temperature changes. Southern Hemisphere countries are in blue; Northern Hemisphere countries are in orange. Thick lines indicate average projected percent change across countries in each hemisphere.

## Discussion

Using a nearly global sample of confirmed COVID-19 cases throughout the course of the recent pandemic, we find that the number of new cases per 1 million individuals responds negatively to rising temperature. This result implies that upcoming seasonal temperature changes will differentially affect COVID-19 transmission across the world. Projecting our estimates onto the next 11 months of expected seasonal temperatures, we predict that countries that will experience colder conditions - primarily in the Southern Hemisphere - are likely to see an increase in COVID-19 transmission during the boreal summer months followed by a drop in transmission during the boreal winter months. Conversely, countries expecting to get warmer during boreal summer months, most of which lie above the equator, will first experience lower COVID-19 transmission before a return to higher rates of transmission. Some important exceptions to this pattern exist, such as in southern India, where an upcoming monsoonal period is likely to lower temperatures from their current levels and raise the rates of transmission, all else equal.

While there is an asymmetric response between Southern and Northern hemisphere countries, there is a clear need for large-scale public health interventions across all countries. For southern countries, which have generally experienced fewer COVID-19 cases to date (Fig. 1A), public health policy must be ramped up in response to rising COVID-19 transmission as temperatures cool. Northern countries face a crucial window of opportunity to eliminate COVID-19 from their populations. In the coming warmer months, rising temperatures are likely to lower overall transmission, making large-scale public health interventions that limit contact amongst individuals, such as shelter-in-place policies and school closings, more effective at achieving and maintaining low overall case rates. However, if such measures are not taken and moderate baseline levels of transmission persist through the summer, cooler fall and winter months could imply a substantial resurgence of new cases (Fig. 2 and 3). Getting the most from these policies during this summer period is critical if northern countries want to eliminate COVID-19, rather than experience a second, possibly stronger, second wave of the virus when temperatures fall during the winter of 2021. History hints at the importance of taking advantage of this window of opportunity: the second wave of the 1918 Spanish influenza pandemic had much higher mortality rates than the first.^22,23^

Although we know of no completed laboratory studies of the temperature-COVID-19 transmission relationship to date, we view our approach as complementary to such future results. While laboratory studies isolate the biology of virus transmission, our statistical approach using observed COVID-19 cases captures those channels *as well as* any behavioral adjustments individuals make in response to short-term fluctuations in temperature, such as decisions to go outside, to exercise, to attend social gatherings, and many other possible activities and health investments. As public health officials grapple with the costs and benefits of a range of possible responses to the current pandemic, quantifying the influence of both channels is essential to building appropriate policies.

Our study has a number of important limitations. First, as is true in any empirical study of disease, we can only observe cases that are confirmed. It is very likely that confirmed cases of COVID-19 fall far below actual rates of infection,^24^ thus suggesting that our findings may represent an under-estimate of the magnitude of the link between infection and local climatic conditions. Relatedly, countries around the world have invested very differently in testing capacity, making such under-reporting heterogeneous across space and time. However, our empirical model is designed to purge estimates of the influence of such differential testing by using a rich set of semi-parametric controls, including fixed effects that vary across both space and time. Second, the estimates we show represent average treatment effects across countries and time periods in our sample. It is possible that the effect of temperature on case rates varies across different policy regimes, such as social distancing and the closure of public transportation systems. While such heterogeneity is an important area for future research, it is important to note that our model, by construction, captures many sources of such heterogeneity through its log-linear form. While we find that temperature has a consistent −13% per 1°C effect on the rate of new cases in a given population, this translates into very different *levels* of cases in different locations. If public health policies successfully lower baseline COVID-19 case rates, changing temperatures will have a much smaller effect on total cases than when baseline rates are high.

## Methods

### Data

#### COVID-19

We use data on global daily new COVID-19 cases assembled by the Johns Hopkins University Center for Systems Science and Engineering^25^ for the period between January 22, 2020 to March 15, 2020.^1^ We aggregate daily new confirmed cases of COVID-19 to the country level, omitting observations from the Diamond Princess cruise ship (due to uncertain temperature exposure of the passengers). Our main outcome variable is the total daily new confirmed cases per 1 million people; we use country-level population in 2018 (the most recent year available) from the World Bank’s World Development Indicators.^2^

#### Weather

We use the ERA5 reanalysis product from European Centre for Medium-Range Weather Forecasts (ECMWF), which provides daily gridded weather variables at the 0.25° latitude by 0.25° longitude resolution.^263^ Specifically, for January 22, 2020 to March 15, 2020, we collect daily mean 2-meter temperature (in degrees centigrade), total 2-meter precipitation (in mm), and mean 1000 hPA specific humidity (in kg/kg). 2-meter and 1000 hPA roughly correspond to conditions near the earth’s surface.

We link gridded climate data to country-level COVID-19 cases by aggregating grid cell information over country boundaries. To preserve any nonlinearities in the temperature-COVID-19 relationship, we take nonlinear transformations of climate variables (e.g. second order polynomials) at the grid cell level before averaging values across space. To capture climatic conditions reflective of population exposure, we average across grid cells weighting by the cross-sectional gridded distribution of population in 2011 from LandScan.^27^ For example, the vector of country-level daily population-weighted average temperature variables we use is computed as 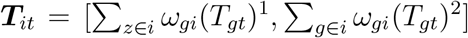, where *g* indicates grid cell, *I* indicates country, superscripts indicate polynomial powers, and *ω*_*gi*_ is the share of country *i*’s population that falls within grid cell *g*.

To estimate projected seasonal temperature conditions and their influence on COVID-19 transmission, we obtain daily gridded 2-meter temperature from ERA5, as described above, over the last five years (2014-2018). Using the average seasonal conditions over the past five years as a proxy for expected seasonal variation through 2020 and into early 2021 (see Fig. S2), we compute monthly averages across all five years of daily temperatures at both grid cell level (see Fig. 2A and C) and at country level (see Fig. 2B and D), in the latter case using the same aggregation method described above.

### Statistical model

We aim to examine whether temperature affects new confirmed cases of COVID-19. In contrast, the goal of most epidemiological models is to explicitly characterize all factors contributing to the transmission of a disease; this endeavor is exceptionally important to guide understanding of disease progression and to forecast total case loads. However, identifying the parameters within such complex models in a plausibly causal manner is difficult. This study takes an econometric approach to isolate random variation in one potentially important driver of transmission, temperature. Our reduced-form empirical approach is agnostic regarding the mechanisms through which temperature influences case rates, but by providing causal estimates of the role that temperature plays in transmission, allows us to make counterfactual simulations of future conditions under alternative temperature conditions. Moreover, these estimates provide empirical grounding for parameterization of other, more structured epidemiological modeling exercises.

There are two challenges to causal estimation in this setting. First, surface temperatures generally fall as one moves away from the equator towards higher latitude locations. Because similar latitude-dependent gradients exist for other potentially relevant environmental conditions like UV exposure and socio-economic indicators like GDP, a cross-sectional analysis of local mean temperature and COVID-19 transmission may be biased by such confounding factors. Second, for a given location, surface temperatures tend to trend over the course of a calendar year. COVID-19 transmission rates trend as well which may also confound temperature effects with other gradually evolving determinants of transmission.

To address these challenges, we conduct a longitudinal (or panel) regression model using daily confirmed COVID-19 cases from 134 countries from January 22, 2020 to March 15, 2020. Our outcome of interest is the total number of new confirmed COVID-19 cases per capita between day *t* and *t −* 1 in country *i*, indicated by Δ*y*_*it*_.^4^ Fig. S1 plots the unconditional distribution of Δ*y*_*it*_ for our country-by-day sample, showing a strongly right-skewed distribution with many zero observations. To address this property of the data, we employ the following Poisson regression model^5^

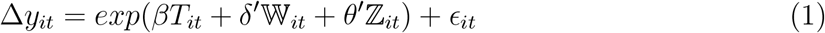

where for country *i* and day *t, T*_*it*_ is population-weighted daily mean temperature (in degrees centigrade). To examine nonlinearity in the relationship between new cases and temperature, in some specifications we also include a quadratic in temperature. 𝕎_*it*_ is a vector of other weather controls, including population-weighted daily total precipitation (in mm) and population-weighted daily specific humidity (in kg/kg). ℤ _*it*_ is a vector of semi-parametric controls designed to isolate random variation in weather conditions.^29^ In our baseline specification, ℤ _*it*_ includes a full set of country-specific dummies, which remove any time-invariant differences in COVID-19 transmission rates across countries. These spatial “fixed effects” address the concern that baseline population characteristics (e.g. economic activity, population density) may be correlated both with transmission and with average temperature. Second, ℤ _*it*_ includes day-specific dummies to remove any common global dynamics in COVID-19 transmission. These temporal fixed effects account for global trends or idiosyncratic shocks in COVID-19 transmission as the disease spreads globally. Finally, in some specifications, ℤ _*it*_ also includes separate quadratic time trends for all countries lying within regularly spaced latitudinal bands (ranging from 5° to 25° wide). These controls, which vary over space and time, remove any potential seasonality in COVID-19 transmission rates that may differ by distance to equator and be correlated with gradually changing seasonal population dynamics, such as time spent outside or work and school schedules. Finally, standard Poisson models impose that the first and second moments of the outcome be equal. To address this overdispersion issue, we cluster standard errors at the country level. This adjustment relaxes the assumption of equal first and second moments by allowing arbitrary forms of within-county heteroskedasticity and serial correlation in the error term ϵ _*it*_.^30^

To conduct our seasonal temperature projections, for each country *i*, we calculate

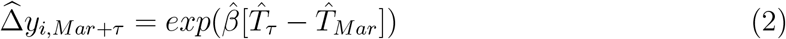

where 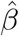 is our estimate from equation (1) (projection values are calculated using the results shown in col. 5 of Table S1). In this ulation 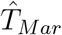 is our constructed estimate of seasonal daily temperature during March, computed as the average daily population-weighted March temperature between 2014-2018. 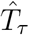 is the seasonal daily temperature during any other month *τ*, also defined as the average daily population-weighted temperature for month *τ* between 2014-2018. In Fig. 2, we show results for *τ* = July (the hottest month on average across the Northern Hemisphere), and *τ* = January (the coldest month on average across the Northern Hemisphere). The estimated impacts, 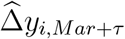 represent the percent change in new COVID-19 cases between March 2020 and future month *τ*, holding all else equal. As described in the main text, these are proportional projections, underscoring the importance of assessing temperature’s influence on COVID-19 alongside the many other factors that determine the baseline case rate at a given location and time.

## Data Availability

All data and replication files can be found at https://github.com/emlab-ucsb/COVID-seasonality.

## Appendix

### Appendix Figures

**Figure S1:**
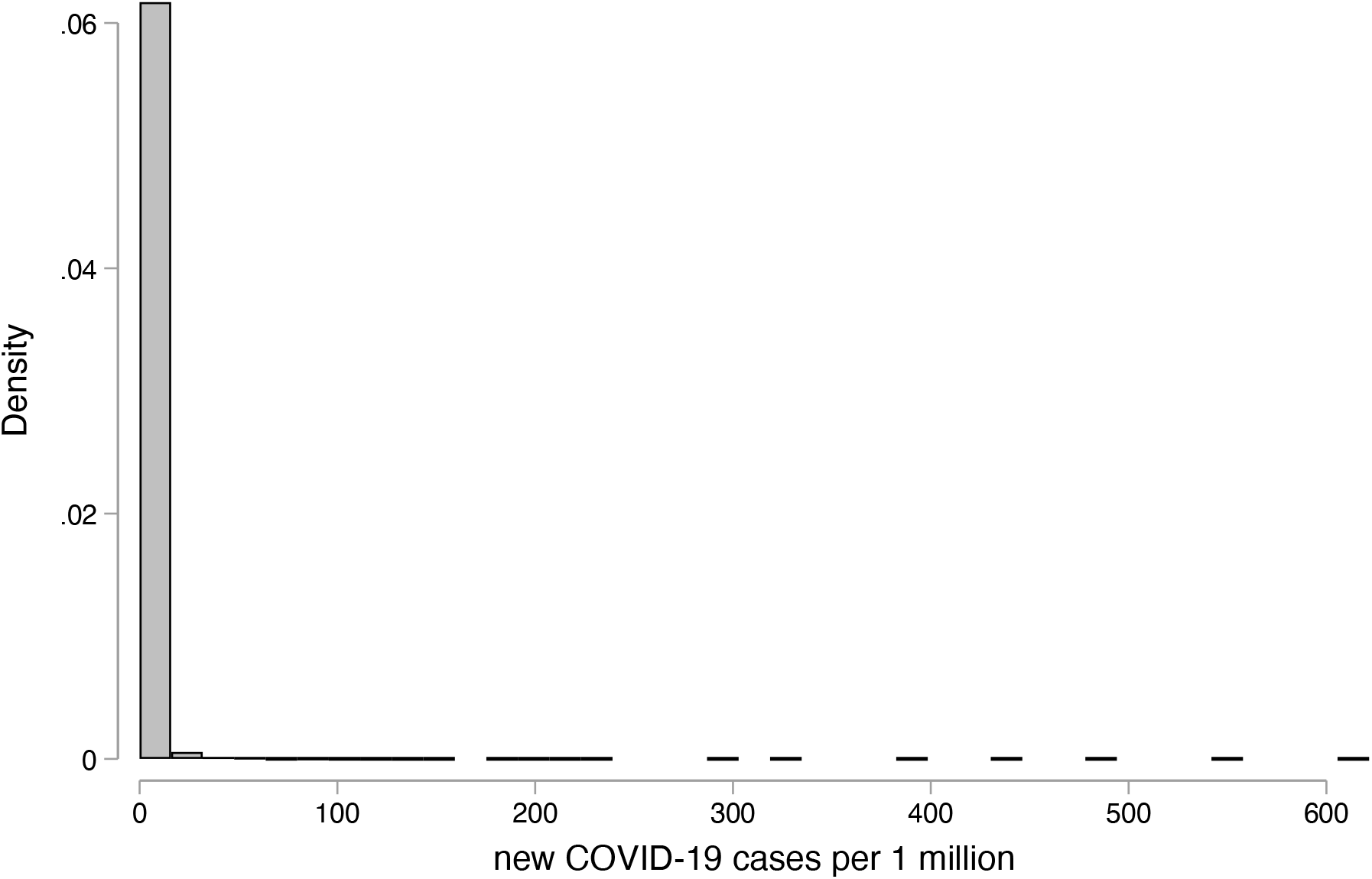
Distribution of sample new COVID-19 cases per 1 million. Figure shows the distribution of new COVID-19 cases per 1 million across country-by-day observations in our estimating sample.

**Figure S2:**
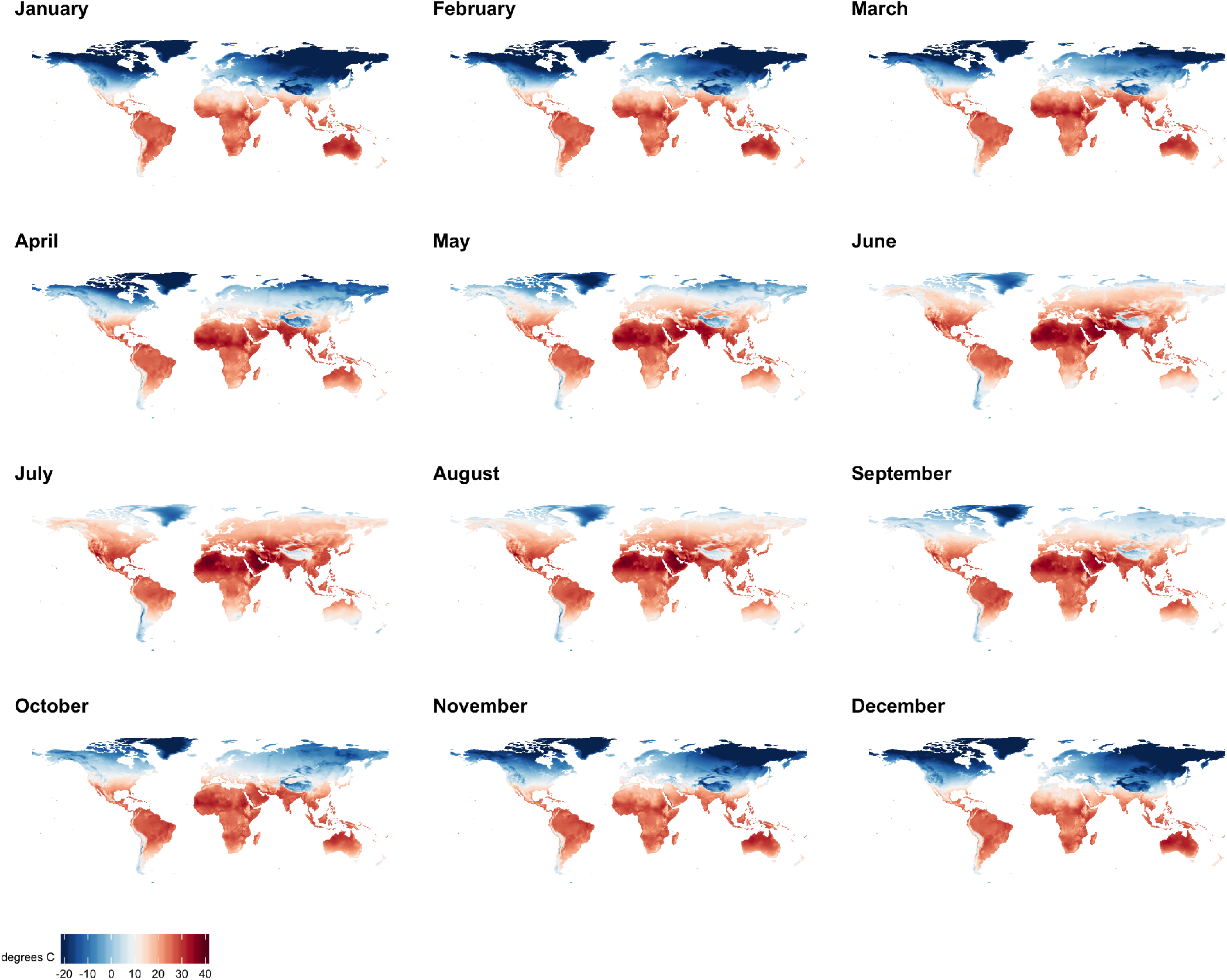
Global gridded monthly temperatures (2014-2018) Panels shows average monthly temperature from January to December at a 0.25° latitude by 0.25° longitude resolution for the 2014-2018 period obtained from the ERA5 reanalysis product generated by the European Centre for Medium-Range Weather Forecasts. These temperatures are used to project seasonal COVID-19 transmission shown in Fig. 2 and 3.

**Figure S3:**
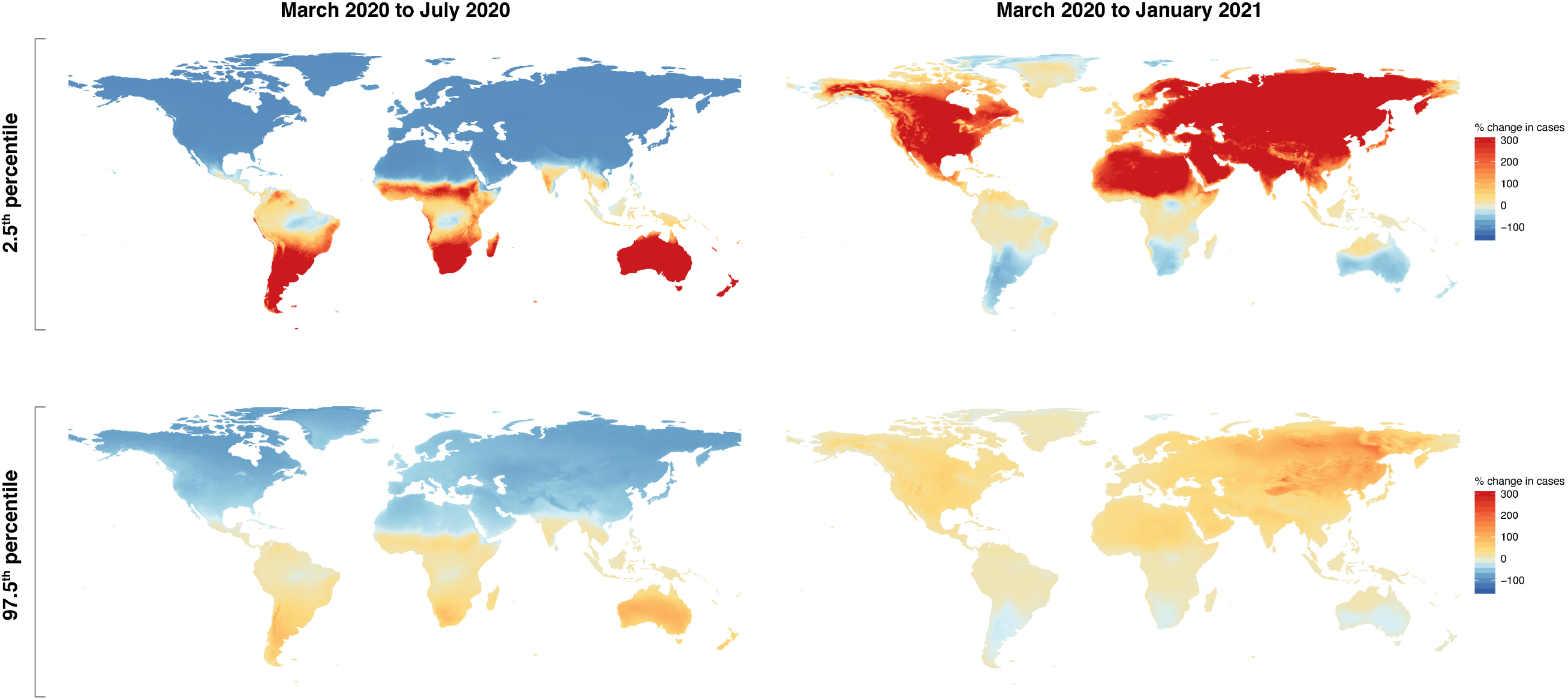
Projected change in COVID-19 cases from March 2020 to July 2020: 2.5th and 97.5th percentiles. Top panel shows the projected percent change in new COVID-19 cases between March 2020 and July 2020 (left) and between March 2020 and January 2021 (right), at a 0.25° latitude by 0.25° longitude resolution using the 2.5 percentile estimates of the linear temperature-COVID-2019 effect, 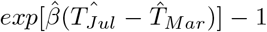, where 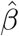 is fromequation (1). Bottom panel replicates the top panel but for the 97.5 percentile estimate of the linear temperature-COVID-19 effect.

### Appendix Tables

**Table S1:** Empirical estimation of the temperature-COVID-19 relationship.

|  | (1) | (2) | (3) | (4) | (5) | (6) | (7) |
| --- | --- | --- | --- | --- | --- | --- | --- |
| Temperature | -0.141***<br>(0.040) | -0.121***<br>(0.043) | -0.111*<br>(0.057) | -0.142***<br>(0.040) | -0.129***<br>(0.043) | -0.127**<br>(0.059) | -0.083*<br>(0.050) |
| Temperature squared |  |  | -0.001<br>(0.003) |  |  | -0.000<br>(0.002) |  |
| Precipitation |  | -0.012<br>(0.017) | -0.013<br>(0.018) |  | -0.023<br>(0.017) | -0.023<br>(0.017) | 0.002<br>(0.014) |
| Specific humidity |  | -37.747<br>(111.012) | -36.518<br>(113.147) |  | -13.998<br>(105.914) | -13.945<br>(106.520) | -73.164<br>(102.965) |
| Observations | 7236 | 7236 | 7236 | 7236 | 7236 | 7236 | 7236 |
| Countries | 134 | 134 | 134 | 134 | 134 | 134 | 134 |
| Days | 54 | 54 | 54 | 54 | 54 | 54 | 54 |
| Weather controls | No | Yes | Yes | No | Yes | Yes | Yes |
| Lat. bin trends | No | No | No | Quad | Quad | Quad | Quad |
| Lat. bin width | - | - | - | 25° | 25° | 25° | 5° |

NOTES: Estimates of Poisson regression model from equation (1) using country-by-day longitudinal data. Outcome is new COVID-19 cases per 1 million population. All models include country and day fixed effects. Column 1 includes linear temperature (in degrees C). Column 2 adds precipitation (in mm) and specific humidity (in kg/kg). Column 3 adds quadratic temperature. Columns 4-6 replicates columns 1-3 but adds separate quadratic time trends for groups of countries within the same 25° latitude bin. Column 7 replicates column 5 but uses a narrower 5° latitude bin. Standard errors clustered at the country-level in parentheses. P-values from two-sided t-tests with \*\*\* $p < 0.01$ , \*\* $p < 0.05$ , \* $p < 0.1$ .

## Footnotes

1 Available at https://github.com/CSSEGISandData/COVID-19 and accessed via https://github.com/RamiKrispin/coronavirus.

2 Available at http://data.worldbank.org/data-catalog/world-development-indicators

3 Available at https://cds.climate.copernicus.eu/cdsapp#!/dataset/reanalysis-era5-single-levels?tab=overview

4 In the canonical S-I-R model of infectious disease transmission, our outcome corresponds to the daily increase in new infected individuals per capita or the daily decrease in susceptible individuals per capita.

5 The main advantage of the Poisson model is that it can accommodate skewed outcome distributions unlike an ordinary least squares model. And unlike log-linear models, it does not drop observations when the outcome variable is zero. The Poisson model has the additional benefit of being a member of the linear exponential family such that even if the outcome variable is not generated by a Poisson process, one can still obtain consistent point estimates through quasi-maximum-likelihood provided that the conditional mean function (i.e., *exp*(*βT*_*it*_ + *δ* ′𝕎 _*it*_ + *θ*′ℤ _*it*_) is ectly specified.^28^

## Notes

### Competing Interest Statement

The authors have declared no competing interest.

### Funding Statement

No external funding was received for this study.

